## Supplementary Materials for "Estimated transmission dynamics of SARS-CoV-2 variants from wastewater are unbiased and robust to differential shedding"

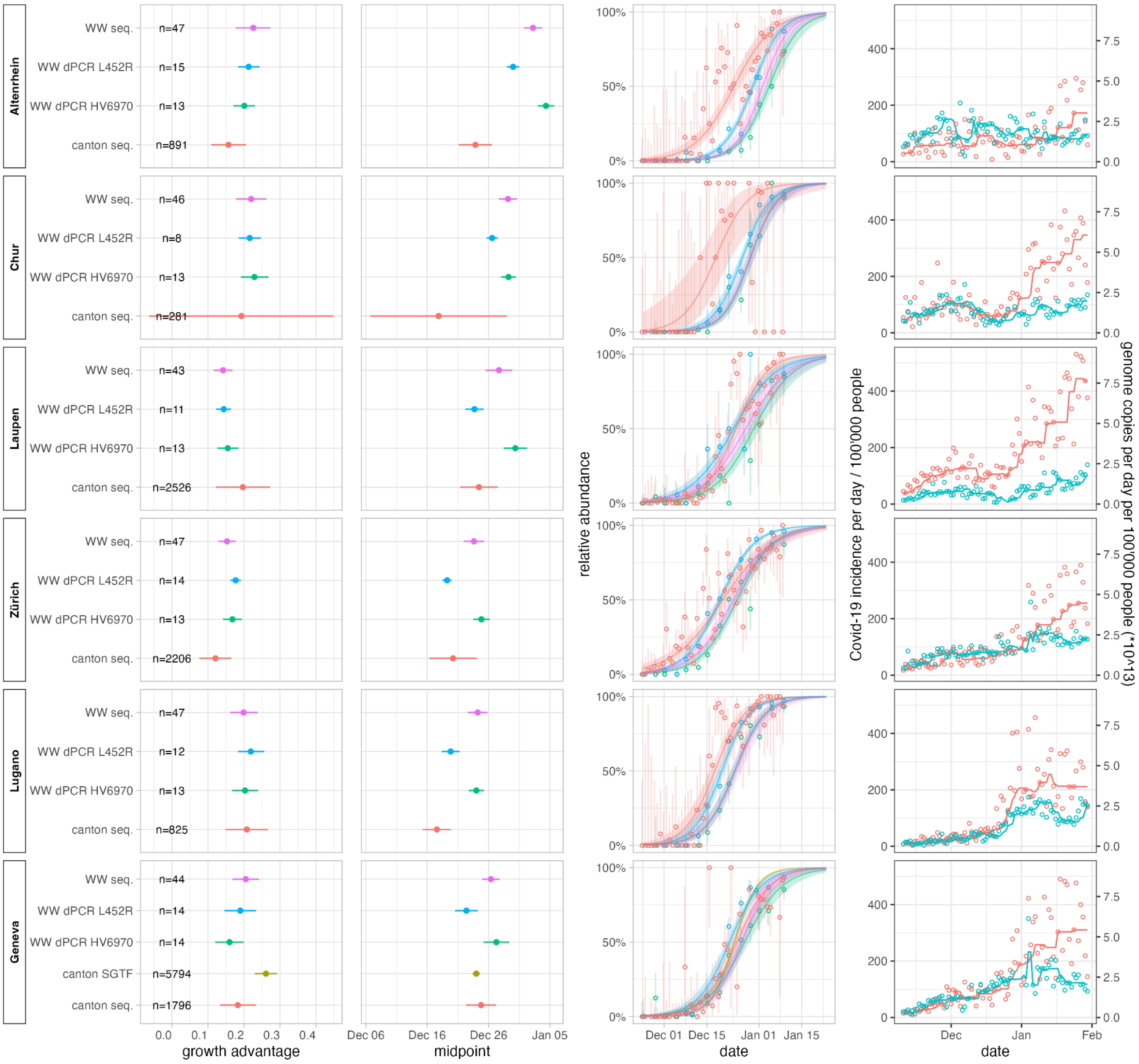


**Supplementary Figure 1:** Concordant estimation of the selection advantage of Omicron BA.1 in Switzerland using clinical and wastewater-derived data. Logistic growth fits of the relative abundance of BA.1 in the six WWTP regions, based on wastewater NGS (WW seq.), wastewater ddPCR duplex assays targeting S:L452R (WW dPCR L452R) and the S:HV69-70 deletion (WW dPCR HV6970), as well as clinical sequencing (canton seq.) and clinical SGTF data (cantong SGTF). Fourth column shows daily new case numbers measured in the surveyed regions, along with measured viral genome copies in wastewater. Third column shows the fitted curves along with 95% Wald confidence bands adjusted for overdispersion, as well as the daily empirical estimates of relative abundance along with 95% confidence intervals. First and second columns respectively show estimates of the selection advantage and midpoint of the logistic growth, along with 95% Wald confidence intervals adjusted for overdispersion.

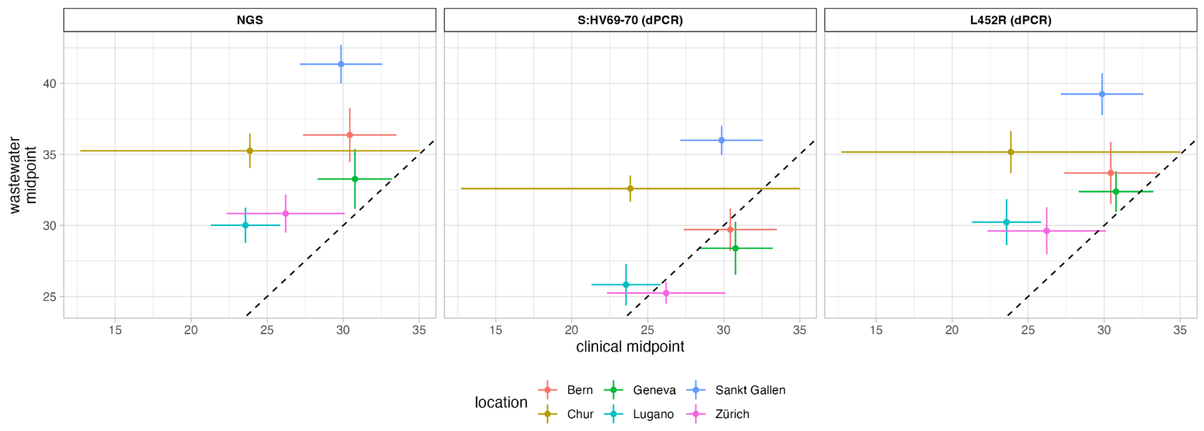


**Supplementary Figure 2:** Midpoint parameter (in days after 2021-11-24) estimates for BA.1 in the six WWTP regions, based on wastewater NGS as well as wastewater ddPCR duplex assays targeting the S:HV69-70 deletion and S:L452R substitution, compared with estimates derived from clinical sequencing from the cantons surrounding the WWTPs. Error bars represent 95% Wald confidence intervals adjusted for overdispersion.

#### Benchmarking S:L452R RT-dPCR Assay

While the Δ69-70 assay was previously designed and evaluated[8], the S:L452R assay has been newly designed for this study. We assessed the performance of the new drop-off RT-dPCR assay against RNA extracts of the SARS-CoV-2 Wuhan-Hu-1 lineage (representing the wild-type) and synthetically assembled RNA fragments (gBlock, Integrated DNA Technologies, USA) representing the VOC following the protocol in Caduff et al. (2022). To assess the performance of the duplex assay estimating the ratio of VOC to wild-type in a mixed sample, we analyzed a series of concentrations containing 0.01, 0.02, 0.10, 0.50, 0.90, 0.98 and 0.99 proportions of VOC RNA (Supplementary Figure 3). All samples were made with molecular grade and contained a total target concentration of 100 gc/μL. Measured proportion of VOC RNA follows the expected proportion, but the ratio of VOC to non-VOC RNA is overestimated by a factor of 1.68 (1.37 – 2.07).

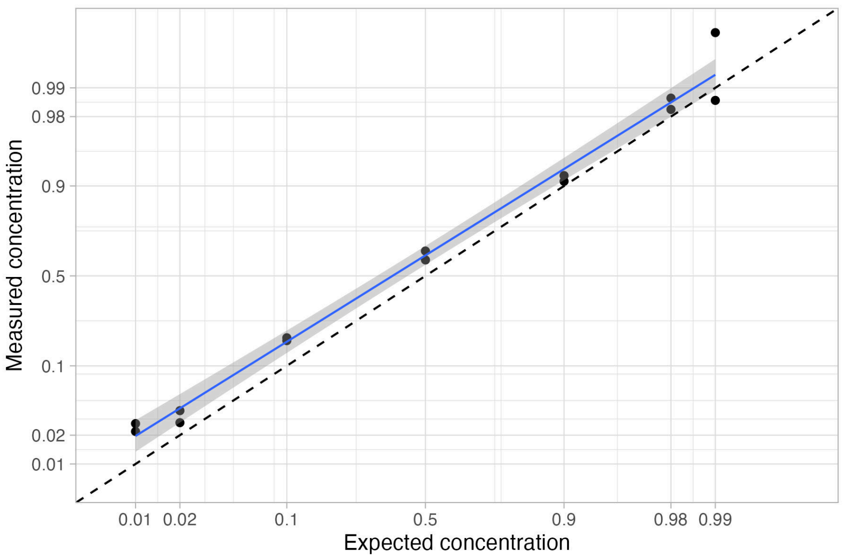

**Supplementary Figure 3:** Benchmarking of the S:L452R drop-off RT-dPCR assay. The relative concentration of RNA from the variant was measured using our newly developed S:L452R RT-dPCR assay in serial dilutions with known relative concentrations. Measured versus expected (from the dilution ratios) proportion of VOC RNA are reported here, both on a logit scale. The plot also shows a linear regression line along with a 95% confidence band. The dotted line shows the 1:1 ideal correspondence. The vertical offset of the linear regression line relative to the dotted line shows that the assay likely overestimates the ratio of variant to non-variant RNA by a constant factor.

**Supplementary Table 1:** Summary of the results, describing shedding related effects on the estimation of selection advantage $s$ of variant $X$ relative to variant $Y$ from wastewater. The first column shows the effect of not accounting for shedding at all, when the shedding load profile (with mean shedding time $\mu$) is the same for both variants. The second column shows the bias $b$ when not accounting for differences in total shedding between variants in the form of a scaling of the shedding load profile by $c$. The third column shows the bias $b$ when not accounting for variant specific shedding load distributions with means $\mu_x$, $\mu_y$ and differences in mean shedding $\Delta_\mu = \mu_y - \mu_x$. We say that the wastewater-based estimates of the selection advantage $\hat{s}^w$ are unbiased if they have no expected difference from the estimate derived from the true relative incidences $\hat{s}$. We say that they are robust if differences in shedding between variants does not introduce any bias.

| | Same shedding profiles | Different total shedding | Different shedding distribution |
|---|---|---|---|
| **Constant growth** | Unbiased | Robust | Robust |
| **Varying growth** | $\hat{s}^w(t) \approx \hat{s}(t - \mu)$ | Robust | $b \approx R_x(t - \mu_y)'\Delta_\mu$ |

**Supplementary Table 2:** Summary of the results, describing shedding related effects on the estimation of the total effective reproduction number $R(t)$ from wastewater. The first column shows the effect of not accounting for shedding at all, when the shedding load profile (with mean shedding time $\mu$) is the same for both variants $X$ and $Y$. The second column shows the bias $b$ when not accounting for differences in total shedding between variants in the form of a scaling of the shedding load profile by $c$. The third column shows the bias $b$ when not accounting for variant specific shedding load distributions with means $\mu_x$, $\mu_y$ and differences in mean shedding $\Delta_\mu = \mu_y - \mu_x$.

| | Same shedding | Different total shedding | Different shedding distribution |
|---|---|---|---|
| **Constant growth** | $R^w(t) \approx R(t - \mu)$ | $b \leq \frac{(c-1)s}{4}$ | $b \leq \frac{s(\beta_x - \gamma)\Delta_\mu}{4}$ |
| **Varying growth** | $R^w(t) \approx R(t - \mu)$ | $b \leq \frac{(c-1)s}{4}$ | $b \leq \frac{s(\beta_x - \gamma)\Delta_\mu}{4} + \Delta_\mu \beta_x'$ |

**Supplementary Table 3. Drop-off RT dPCR assay primer and probe sequences.**

| Target | Type | DNA oligo name | DNA Sequence (5’-3’) | Amplicon size | Reference |
|---|---|---|---|---|---|
| *Delta* | *Primer forward* | *L452R_delta_F* | *TGATAGATTTCAGTTGAAATATCTCTCTCA* | | *Current study* |
| | *Primer reverse* | *L452R_delta_R* | *AATCTTGATTCTAAGGTTGGTGGTAATTAT* | | |
| | *Probe mutation* | *L452R_delta_P_mut* | *CTAAACAATCTATACCGGTAAT* | | |
| | *Probe Wildtype* | *L452R_delta_P_wtcomp* | *CCTAAACAATCTATACAGGTAA* | | |
| *Alpha/Omicron* | *Primer forward* | *Yale_69-70_F* | *TCAACTCAGGACTTGTTCTTACCT* | | *Vogels et al. 2021, Caduff et al. 2022* |
| | *Primer reverse* | *Yale_69-70_R* | *TGGTAGGACAGGGTTATCAAAC* | | |
| | *Probe Wildtype* | *Yale_69-70_Cy5_P* | *TTCCATGCTATACATGTCTCTGGGA* | | |
| | *Probe mutation* | *LC_69-70_HEX_P* | *CCAATGGTACTAAGAG* | | *Caduff et al., 2022* |

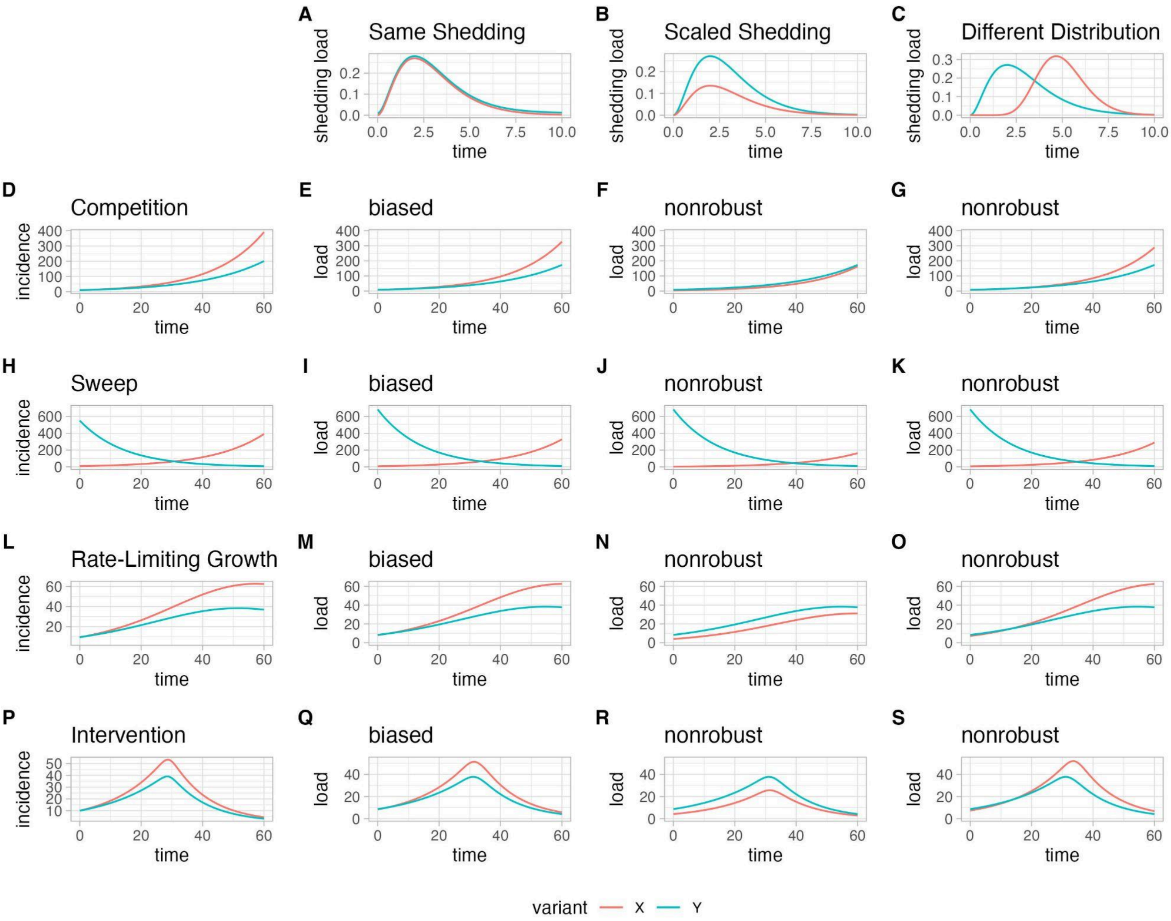


**Supplementary Figure 4**: convolution of different time series of incidence of variants $X$ and $Y$ (rows) with different shedding load profiles for variants $X$ and $Y$ (columns). We show 3 scenarios for the shedding: same shedding profiles for both variants (**A**), 50% lower total shedding for $X$ relative to $Y$ (**B**), and shedding load distribution for $X$ with arbitrary different shape relative to $Y$ (but with the same total shedding, **C**). Plots at the row index show 4 different scenarios for the dynamics of the incidence of variants $X$ and $Y$: the first row (**D**), two variants with constant (but different) transmission rates are introduced at the same time ; second row (**H**), two variants with constant (but different) transmission rates, one is replacing the other ; third row (**L**), two variants are introduced at the same time, their transmission rates decreasing smoothly according to the rate-limiting mass action dynamics of a SIR model ; fourth row (**P**), two variants are introduced at the same time, their transmission rates are abruptly cut to almost zero by an intervention in the middle of the time series. After being calibrated, the estimates of incidence are only moderately biased with respect to the shedding of variants, generally in the form of a shift and smoothing (**E**, **I**, **M**, **Q**). Differences in total shedding decouple break the calibration and introduce a large error (**F**, **J**, **N**, **R**). Different shedding dynamics introduce a mild error (**G**, **K**, **O**, **S**).

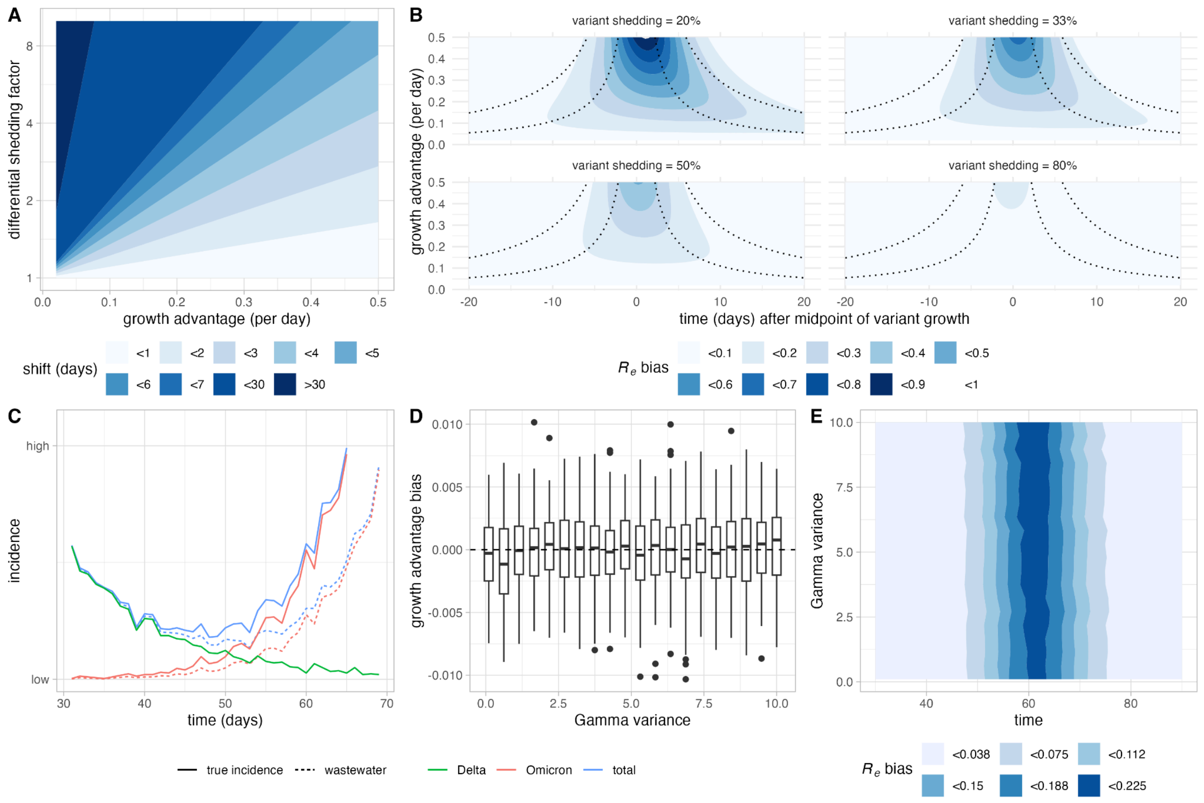


**Supplementary Figure 5:** Dependence of the bias in midpoint and $R_e$ estimation on shedding difference, selection advantage, and generation interval time distribution. **A:** The time-shift in the growth and decay curves depends both on the magnitude of differential shedding and on the selection advantage of the variant. **B:** The magnitude of the bias of the $R_e$ estimate depends on the amount of undershedding. The bias rises through time and vanishes as the variant replaces the other strain. For variants with a higher selection advantage, the bias rises and decreases more sharply. Dotted lines indicate when the variant reaches 5%, 25%, 75%, and 95% of infections (left to right). **C:** Simulated time series, where a variant with $R_e$=0.6 (Delta, green) is replaced by a newly introduced variant with $R_e$=2.2 (Omicron, red). The generation time is sampled from a Gamma distribution with mean 4.8 days and variance 5 days$^2$. The new variant is assumed to shed 50% less, leading to underestimation of its incidence (solid line) from wastewater concentrations (dashed line) and underestimation of the total incidence of the virus (blue). **D:** For increasing variance levels of the generation interval time distribution (20 values linearly spaced between 0.1 and 10.0 days, but constant mean of 4.8 days), we produced selection advantage estimates from simulated data with and without undershedding and calculated the difference to estimate the bias stemming from undershedding. At each variance level, we repeated the simulation 100 times. Boxplots hinges represent the median and quartiles, with whiskers extending to the largest and smallest values no further than 1.5 times the interquartile range (data points beyond these limits are plotted individually). On average, the bias is zero, independently of the variance of the generation interval time distribution. **E:** Using the same simulations as in B, the bias in $R_e$ over time averaged over the simulation runs is shown. The transient pattern of the bias is not affected by the variance of the generation interval time distribution.

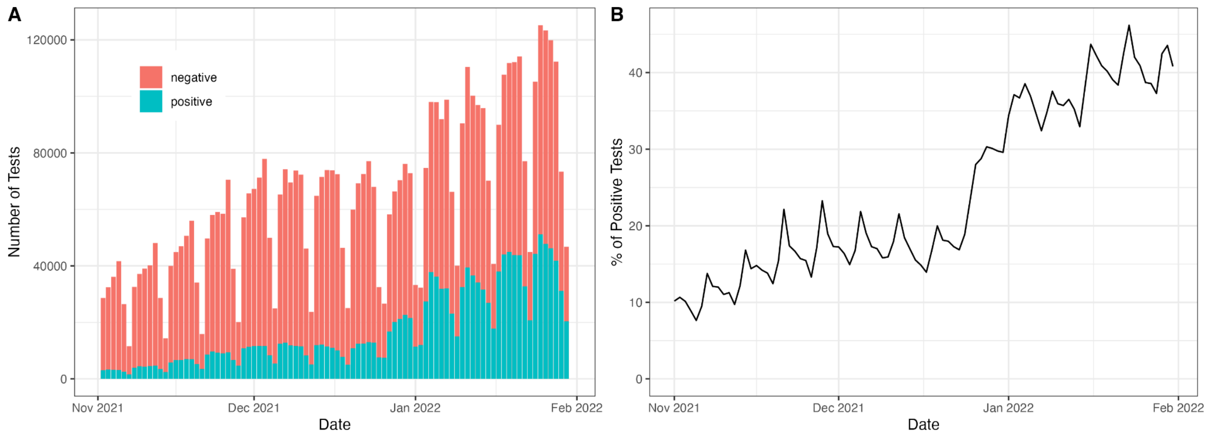


**Supplementary Figure 6:** Number of tests and test positivity rate in Switzerland. **A:** The number of both negative and positive tests increased during the period when Omicron BA.1 spread in Switzerland. **B:** The test positivity rate (number of positive tests divided by the total number of tests) increased during the spread of BA.1 in Switzerland, indicating likely increase in the underreporting of positive cases. Data recovered from corona-data.ch.

### A. Model of competition between variants

We describe here the model describing the competition between two variants $X$ and $Y$ in the population. We assume that the population is large and reproducing in continuous time. The incidence of variants $X$ and $Y$ varies following the ODE system

$$X'(t) = \beta_x X(t) - \gamma X(t) \text{ and } Y'(t) = \beta_y Y(t) - \gamma Y(t) \quad (1)$$

In this model, $\beta$ and $\gamma$ represent the transmission and recovery rates (i.e., the rate at which infected individuals infect non-infected individuals, and the rate at which they themselves become uninfected), respectively. In general, $\beta$ and $\gamma$ are assumed to be constant through time, with $\gamma$ equal for all variants. The reproduction number $R$ is given by

$$R = \frac{\beta}{\gamma} \quad (2)$$

Because $\beta$ is assumed to be constant for both variants, it follows that their incidence grows (or decays) exponentially

$$X(t) = X(0)\exp\{(\beta_x - \gamma)t\} \text{ and } Y(t) = Y(0)\exp\{(\beta_y - \gamma)t\} \quad (3)$$

Where $X(0)$ and $Y(0)$ are the incidence of the variants at time $t = 0$. The relative abundance $f$ of variant $X$ follows the logistic growth

$$f(t) = \frac{X(t)}{X(t) + Y(t)} = \frac{\exp\{(\beta_x - \beta_y)(t - t_0)\}}{1 + \exp\{(\beta_x - \beta_y)(t - t_0)\}} \quad (4)$$

where $t_0$ is the midpoint of the curve where both variants are equally abundant. For the derivative on the logit-transformed relative abundances $\phi(t) = \log(\frac{f(t)}{1-f(t)}) = \log(\frac{X(t)}{Y(t)})$, we find

$$\phi' = X(t)^{-1}X'(t) - Y(t)^{-1}Y'(t) = \beta_x - \beta_y \quad (5)$$

Defining the selection advantage of variant $X$ over variant $Y$ as $s = \beta_x/\beta_y - 1$, such that $R_x/R_y = 1 + s$, we obtain

$$\phi'\gamma^{-1} = sR_Y \quad (6)$$

Thus, to estimate $s$ from a time series of measurements of $f(t)$, we estimate $\phi'$, multiply it by the (assumed known) average generation time $\gamma^{-1}$, and assume $R_Y \approx 1$. We will denote this estimate by $\phi'\gamma^{-1} = \hat{s}$. Generally, $\phi'$ can be readily estimated as the rate parameter of logistic regression of $f(t)$.

##### Non-constant infection rates

When dropping the assumption of constant infection rates, $\phi'(t)$ is not necessarily constant through time anymore,

$$\phi'(t) = \beta_x(t) - \beta_y(t) \quad (7)$$

This is particularly true when maintaining the constraint of a constant selective advantage $s = \beta_x(t)/\beta_y(t) - 1$. In this case, instantaneous estimates of $\phi'(t)\gamma^{-1} = sR_Y(t)$, will yield what we will refer to here as the apparent selective advantage, independent of the procedure used to obtain them. The infection rates $\beta_x(t)$, $\beta_y(t)$ could vary for a multitude of reasons, including behavioral changes in the population. An interesting class of models in which they are not constant is in so-called mass action type models, such as the SIR model. In those models, the growth rates are limited by the currently available pool of susceptible individuals $S(t)$, or more precisely by their fraction relative to the population size $N$ (assumed constant)

$$X'(t) = \beta_x \frac{S(t)}{N} X(t) - \gamma X(t) \text{ and } Y'(t) = \beta_y \frac{S(t)}{N} Y(t) - \gamma Y(t) \quad (8)$$

#### B. Convolution with the shedding load profile

We do not directly observe the incidence of $X$ and $Y$. Instead, the viral loads measurable in a wastewater sample correspond to the incidences convolved with the shedding load profile (SLP) $g$, which represents the average shedding of viral particles through time after infection. We thus have the following form of the loads $X^w$ and $Y^w$ of both variants

$$X^w(t) = (X * g)(t) = \int_\tau X(\tau)g(t - \tau)d\tau \text{ and } Y^w(t) = (Y * g)(t) = \int_\tau Y(\tau)g(t - \tau)d\tau \quad (9)$$

The observed loads will generally track the incidence through time, although the convolution with the SLP will have a shifting and smoothing effect (Supplementary Figure 4). We can approximate this bias as follow: We consider a second-degree Taylor approximation of $X(t)$ such that

$$X^w(t) \approx \int_\tau \left(X(t) - X'(t)\tau - X''(t)\frac{\tau^2}{2}\right)g(\tau)d\tau \quad (10)$$

$$= X(t)\int_\tau g(\tau)d\tau - X'(t)\int_\tau g(\tau)\tau d\tau - X''(t)\frac{1}{2}\int_\tau g(\tau)\tau^2 d\tau$$

$$= l\left(X(t) - X'(t)\mu - X''(t)\frac{\mu^2}{2} - X''(t)\frac{\sigma^2}{2}\right)$$

$$\approx l\left(X(t - \mu) - X''(t)\frac{\sigma^2}{2}\right)$$

where $l = \int_\tau g(\tau)d\tau$ is the expected total amount of viral particles shed through the course of infection, $\mu = E\left[g(\tau)/l\right]$ and $\sigma^2 = Var\left[g(\tau)/l\right]$ are the expected value and variance of the normalized shedding load profiles, respectively. We thus see that the relationship between the observed viral loads in wastewater and the incidence of the variant is as follows: the observed viral loads are scaled by l, shifted in time by $-\mu$, and the peaks and troughs are smoothed proportionally to $\sigma^2$. Similarly, a first-degree Taylor expansion leads to the approximation

$$X^w(t) \approx l\, X(t - \mu) \quad (11)$$

### C. Bias in the estimates of selection advantage and robustness to changes in shedding load profile

In this section, we turn our attention to biases in the estimation of the selection advantage from relative load data. We say that the estimates are unbiased when they are the same as the ones that are obtained from the actual relative incidence, and we say they are robust if changes in the shedding load profile from a variant to another do not add or remove bias.

#### (i) Estimates are unbiased and robust under constant infection rates

First, assuming continuity and compact support, we apply Leibniz integral rule

$$\frac{d}{dt}X^{w}(t) = \frac{d}{dt}(X * g)(t) = \int_{\tau} \frac{\partial}{\partial t}X(t - \tau)g(\tau)d\tau = (X' * g)(t) \quad (12)$$

and similarly

$$\frac{d}{dt}Y^{w}(t) = (Y' * g)(t) \quad (13)$$

Now, assuming constant infection rates, and by using commutativity of the convolution product with scalar multiplication

$$\frac{d}{dt}X^{w}(t) = ((\beta_x - \gamma)X * g)(t) = (\beta_x - \gamma)(X * g)(t) \quad (14)$$

and similarly

$$\frac{d}{dt}Y^{w}(t) = (\beta_y - \gamma)(Y * g)(t) \quad (15)$$

From the observed relative abundance of loads $f^{w}$ and its logit-transformed values $\phi^{w}$, we obtain

$$\frac{d}{dt}\phi^{w}(t) = X^{w}(t)^{-1}\frac{d}{dt}X^{w}(t) - Y^{w}(t)^{-1}\frac{d}{dt}Y^{w}(t) \quad (16)$$
$$= (\beta_x - \gamma)(X * g)(t)(X * g)(t)^{-1} - (\beta_y - \gamma)(Y * g)(t)(Y * g)(t)^{-1} = \beta_x - \beta_y$$

And therefore the estimates of selective advantage from wastewater data $\hat{s}^{w}$ are unbiased

$$\hat{s}^{w} = \frac{d}{dt}\phi^{w}(t)\gamma^{-1} = (\beta_x - \beta_y)\gamma^{-1} = \phi'\gamma^{-1} = \hat{s} \quad (17)$$

Importantly, arbitrarily changing the SLP for the variant $X$ from $g$ to an arbitrary $g_x$ has no impact on the above results:

$$\frac{d}{dt}\phi^{w_2}(t) = (X * g_x)(t)^{-1}\frac{d}{dt}(X * g_x)(t) - (Y * g)(t)^{-1}\frac{d}{dt}(Y * g)(t) = \beta_x - \beta_y \quad (18)$$

#### (ii) Estimates are always robust to changes in total shedding

Next, we show that the estimates do not incur any bias when the total amount of shedding changes, even when infection rates are not constant or when the selective advantage $s$ is not constant. We assume that $X$ is shed according to a scaled shedding load profile $g_s(t) = cg(t)$. By commutativity of convolution and scalar multiplication, we have

$$X^{w_s}(t) = (X * g_s)(t) = (X * cg)(t) = c(X * g)(t) \quad (19)$$

From the observed relative abundance of loads $f^{w_s}$ and its logit-transformed values $\phi^{w_s}$ we obtain

$$\frac{d}{dt}\phi^{w_s}(t) = (c(X * g)(t))^{-1}\frac{d}{dt}(c(X * g)(t)) - (Y * g)(t)^{-1}\frac{d}{dt}(Y * g)(t) \quad (20)$$
$$= (X * g)(t)^{-1}\frac{d}{dt}(X * g)(t) - (Y * g)(t)^{-1}\frac{d}{dt}(Y * g)(t)$$

which does not depend on $c$.

However, changes in total shedding will lead to bias in estimation of the relative incidence of variants,

$$\phi^{w_s}(t) = \log\{(X * cg)(t)\} - \log\{(Y * g)(t)\} = \phi^{w}(t) + \log(c) \quad (21)$$

Thus, the logit-transformed estimates of wastewater-based relative incidences are shifted by a constant $\log(c)$. If the standard assumption of constant transmission rates holds and hence $f(t)$ follows logistic growth, then estimates of the midpoint $t_0$ where $f(t_0) = 0.5$ (i.e., $X$ amounts for 50% of the incidence) will incur a bias (Supplementary Figure 5)

$$t_0^{w_2} = t_0^{w} - \log(c)/s \quad (22)$$

#### (iii) Estimates are approximately shifted when dropping the constant infection rates assumption

The common methods to estimate the selective advantage assume that the infection rates are constant through time. We have shown above that when this assumption does not hold, the instantaneous estimates of the selective advantage, computed from true relative incidence, will vary through time. We referred to those as apparent selection advantages. We now investigate what happens to those apparent selective advantages when computed from relative wastewater loads instead of true relative incidence, in the case of non-constant infection rates. We thus have the incidences of $X$ and $Y$ evolving as

$$X'(t) = \beta_x(t)X(t) - \gamma X(t) \text{ and } Y'(t) = \beta_y(t)Y(t) - \gamma Y(t) \quad (23)$$

with $\beta_x(t)$ and $\beta_y(t)$ not constant. We now look at the time derivative of the logit relative abundance, and make first order Taylor approximations of $X(t)$ and $Y(t)$:

$$\frac{d}{dt}\phi^{w}(t) = \frac{(X*g)'(t)}{(X*g)(t)} - \frac{(Y*g)'(t)}{(Y*g)(t)} \approx \frac{\frac{d}{dt}\int_\tau (X(t)-X'(t)\tau)g(\tau)d\tau}{\int_\tau (X(t)-X'(t)\tau)g(\tau)d\tau} - \frac{\frac{d}{dt}\int_\tau (Y(t)-Y'(t)\tau)g(\tau)d\tau}{\int_\tau (Y(t)-Y'(t)\tau)g(\tau)d\tau} \quad (24)$$

$$=\frac{\frac{d}{dt}\{X(t)-X'(t)\mu\}}{X(t)-X'(t)\mu} - \frac{\frac{d}{dt}\{Y(t)-Y'(t)\mu\}}{Y(t)-Y'(t)\mu}$$

$$\approx \frac{\frac{d}{dt}\{X(t-\mu)\}}{X(t-\mu)} - \frac{\frac{d}{dt}\{Y(t-\mu)\}}{Y(t-\mu)}$$

$$= \beta_x(t - \mu) - \beta_y(t - \mu) = s\beta_y(t - \mu)$$

The apparent selective advantage is shifted in time by $\mu$ relative to the one obtained from the relative incidence

$$\hat{s}^w(t) \approx s\beta_y(t - \mu)\gamma^{-1} = \hat{s}(t - \mu) \quad (25)$$

Thus, at any given time, the estimates will have an offset relative to those computed from true relative incidences approximately proportional to $\beta'_y(t)$ and $\mu$

$$\hat{s}^w(t) - \hat{s}(t) \approx \hat{s}(t - \mu) - \hat{s}(t) \approx \hat{s}(t) - \hat{s}'(t)\mu - \hat{s}(t) =- \beta'_y(t)\mu s\gamma^{-1} \quad (26)$$

And the relative bias will be approximately proportional to $\beta'_y(t)/\beta_y(t)$ and $\mu$,

$$\frac{\hat{s}^w(t)-\hat{s}(t)}{\hat{s}(t)} \approx -\frac{\beta'_y(t)}{\beta_y(t)}\mu \quad (27)$$

#### (iv) Estimates are approximately shifted when dropping the constant selection assumption

Most models of selection assume a fixed, constant multiplicative selection advantage $s$. If that assumption is relaxed, we have $\beta_x(t)$ not constant, and from above

$$\frac{d}{dt}\phi^w(t) \approx \beta_x(t - \mu) - \beta_y(t - \mu) \quad (28)$$

We assume without loss of generality that $\beta_y$ is constant, such that

$$\frac{d}{dt}\phi^w(t) \approx \beta_x(t - \mu) - \beta_y = (1 + s(t - \mu))\beta_y - \beta_y \quad (29)$$

Similarly as above, the apparent selection advantage are shifted in time by $\mu_g$ relative to the ones from the true relative incidences

$$\hat{s}^w(t) \approx s(t - \mu)\gamma^{-1}\beta_y = \hat{s}(t - \mu) \quad (30)$$

Following the same argument as above, these estimates will be robust to changes in total shedding for variant $X$.

#### (v) Estimates are unbiased and robust to non-constant generation times

Models of selection generally assume constant generation time $\gamma^{-1}$. If we relax this assumption, such that

$$X'(t) = \beta_x X(t) - \gamma(t)X(t) \text{ and } Y'(t) = \beta_y Y(t) - \gamma(t)Y(t) \tag{31}$$

we find

$$\frac{d}{dt}\phi^{w}(t) \approx \frac{\frac{d}{dt}\{X(t-\mu)\}}{X(t-\mu)} - \frac{\frac{d}{dt}\{Y(t-\mu)\}}{Y(t-\mu)} = \beta_x - \beta_y - \gamma(t-\mu) + \gamma(t-\mu) = \beta_x - \beta_y \tag{32}$$

If we now use a fixed generation time $\gamma^{-1}$ to translate $\phi^{w}(t)$' into an estimate of the selection, we will indeed incur an error. That error is independent of shedding such that the estimates of selection based on wastewater relative loads are equal to those based on the true relative incidence, i.e. they are unbiased and robust w.r.t shedding.

#### (vi) Estimates are non-robust under non-constant infection rates and arbitrary transformations of the shedding load profile

If we now assume that $X$ and $Y$ have different shedding load distributions $g_x$ and $g_y$ with expected values $\mu_x$ and $\mu_y$ such that $\Delta_\mu = \mu_x - \mu_y$, then

$$\begin{aligned} \frac{d}{dt}\phi^{w_2}(t) &\approx \beta_x(t-\mu_x) - \beta_y(t-\mu_y) \\ &= (1+s)\beta_y(t-\mu_y-\Delta_\mu) - \beta_y(t-\mu_y) \\ &\approx (1+s)\beta_y(t-\mu_y) - (1+s)\beta'_y(t-\mu_y)\Delta_\mu - \beta_y(t-\mu_y) \\ &= s\beta_y(t-\mu_y) - (1+s)\beta'_y(t-\mu_y)\Delta_\mu \end{aligned} \tag{33}$$

such that the bias will be approximately proportional to $\beta'_y(t-\mu_y)$ and $\Delta_\mu$,

$$\hat{s}^{w_2}(t) - \hat{s}^{w}(t) \approx (1+s)\beta'_y(t-\mu_y)\Delta_\mu\gamma^{-1} = R_x(t-\mu_y)'\Delta_\mu \tag{34}$$

and the relative bias will be approximately proportional to $\beta'_y(t-\mu_y)/\beta_y(t-\mu_y)$ and $\Delta_\mu$,

$$\frac{\hat{s}^{w_2}(t) - \hat{s}^{w}(t)}{\hat{s}^{w}(t)} \approx -\frac{\beta'_y(t-\mu_y)}{\beta_y(t-\mu_y)}\Delta_\mu\frac{1+s}{s} \tag{35}$$

#### (vii) Estimates are unbiased and robust to changes in generation time

Next, we consider a model in which not only the infection rates but also the generation times are different between variants. The infection rates can be assumed time-dependent, while we will assume that the generation times are fixed,

$$X'(t) = \beta_x(t)X(t) - \gamma_x X(t) \ and \ Y'(t) = \beta_y(t)Y(t) - \gamma_y Y(t) \quad (36)$$

When computed from true relative incidence, the apparent selection advantage will be biased if similar generation times $\gamma_y^{-1}$ are assumed for both variants

$$\phi'(t) = \beta_x(t) - \beta_y(t) - \left(\gamma_x - \gamma_y\right) \Rightarrow \hat{s}(t) = \phi'(t)\gamma_y^{-1} = sR_y(t) - \frac{\gamma_x - \gamma_y}{\gamma_y} \quad (37)$$

In the case of estimates based on the observed wastewater loads, we note that

$$\begin{aligned}(X * g_x)'(t) &= \int_\tau X'(t-\tau)g_x(\tau)d\tau = \int_\tau \left(\beta_x(t-\tau) - \gamma_x\right)X(t-\tau)g_x(\tau)d\tau \\ &= (\beta_x X * g_x)(t) - \gamma_x(X * g_x)(t)\end{aligned} \quad (38)$$

and similarly

$$(Y * g)'(t) = (\beta_y Y * g)(t) - \gamma_y(Y * g)(t) \quad (39)$$

The expression for an eventual bias is then

$$\begin{aligned}\frac{d}{dt}\phi^{w_2}(t) - \phi'(t) &= \left(\frac{(X*g_x)'(t)}{(X*g_x)(t)} - \frac{(Y*g)'(t)}{(Y*g)(t)}\right) - \left(\beta_x(t) - \beta_y(t) - \left(\gamma_x - \gamma_y\right)\right) \\ &= \left(\frac{(\beta_x X*g_x)(t)}{(X*g_x)(t)} - \frac{(\beta_y Y*g)(t)}{(Y*g)(t)} - \left(\gamma_x - \gamma_y\right)\right) - \left(\beta_x(t) - \beta_y(t) - \left(\gamma_x - \gamma_y\right)\right) \\ &= \left(\frac{(\beta_x X*g_x)(t)}{(X*g_x)(t)} - \frac{(\beta_y Y*g)(t)}{(Y*g)(t)}\right) - \left(\beta_x(t) - \beta_y(t)\right)\end{aligned} \quad (40)$$

The bias does not depend on $\left(\gamma_x - \gamma_y\right)$, hence the estimates are unbiased and robust.

#### (viii) Bias and robustness of estimates to variant-specific generation times

In the case where we additionally have non-constant generation times, such that

$$X'(t) = \beta_x X(t) - \gamma_x(t)X(t) \ \text{and} \ Y'(t) = \beta_y Y(t) - \gamma_y(t)Y(t) \quad (41)$$

the estimates of selection based on true relative incidence will incur now a bias non-constant in time:

$$\hat{s}(t) = \gamma^{-1}\left(\left(\beta_x - \beta_y\right) - \left(\gamma_x(t) - \gamma_y(t)\right)\right) \quad (42)$$

where $\gamma^{-1}$ is the generation time. Following the Taylor approximations of $X(t)$ and $Y(t)$ described above, we have for the estimates based on relative loads,

$$\hat{s}^w(t) = \gamma^{-1}\left(\left(\beta_x - \beta_y\right) - \left(\gamma_x(t - \mu) - \gamma_y(t - \mu)\right)\right) \approx \hat{s}(t - \mu) \quad (43)$$

i.e, the bias is shifted in time, and the estimates remain robust to differences in total shedding.

If in addition, we now assume that $\gamma_x^{-1}$ and $\gamma_y^{-1}$ are variant-specific but do not vary over time, then we have that:

$$\hat{s}^w = \gamma^{-1}\left(\left(\beta_x - \beta_y\right) - \left(\gamma_x - \gamma_y\right)\right) = \hat{s} \quad (44)$$

I.e. in that case, $\hat{s}^w$ is both unbiased and robust with respect to shedding, despite the variant-specific generation times.

### D. Bias in $R_e$ estimates

Next, we analyze the effective reproduction number $R_e$. We derive a closed-form approximation for the bias in $R_e$ estimation due to lower shedding of a variant and show that it is nonzero for a short period of time. We provide an upper bound for the bias.

Let $I(t) = X(t) + Y(t)$ be the total incidence through time. Following Supplementary Equation 2, we have

$$R(t) = \beta(t)\gamma^{-1} = 1 + \gamma^{-1}\log(I(t))' = 1 + \gamma^{-1}\frac{I(t)'}{I(t)} \quad (45)$$

#### (i) Estimates are shifted when not accounting for shedding

Using the approximation from Supplementary Equation 11, convolving the incidence $I(t)$ with shedding load profile $g(t)$ yields a biased estimator for the reproduction number

$$R^w(t) = 1 + \gamma^{-1}\frac{(I*g)(t)'}{(I*g)(t)} \approx 1 + \gamma^{-1}\frac{lI(t-\mu)'}{lI(t-\mu)} = R(t - \mu) \quad (46)$$

Importantly, the total loads here cancel out, such that the shape of the shedding load distribution (and mainly its first moment) matter only. Methods to estimate the reproduction number from wastewater typically assume that the shedding load profile (or at least, distribution) is known, and deconvolve the loads first before estimation of $R_e$, such that

$$R^{w_1}(t) = 1 + \gamma^{-1}\frac{(I*g*g^{-1})(t)'}{(I*g*g^{-1})(t)} = 1 + \gamma^{-1}\frac{I(t)'}{I(t)} = R(t) \quad (47)$$

#### (ii) Estimates are biased by differences in total shedding

Now let's assume we wrongly assume that $g_x = g_y$, while in fact total shedding is different such that $g_x = cg_y$. We then have the bias for our estimator $R^{w_2}(t)$

$$R^{w_2}(t) - R(t) = 1 + \gamma^{-1}\log\left(\left(X * g_x + Y * g_y\right) * g_y^{-1}\right)' - \left[1 + \gamma^{-1}\log\left(X(t) + Y(t)\right)'\right] \quad (48)$$

$$= \gamma^{-1}\log\left(cX(t) + Y(t)\right)' - \gamma^{-1}\log\left(X(t) + Y(t)\right)'$$

$$= \gamma^{-1}\log\left(\frac{cX(t)+Y(t)}{X(t)+Y(t)}\right)' = \gamma^{-1}\log\left(cf(t) + (1 - f(t))\right)'$$

$$= \gamma^{-1}\log\left(1 + (c - 1)f(t)\right)' = \gamma^{-1}\frac{(c-1)f(t)'}{1+(c-1)f(t)}$$

Importantly this shows that the bias depends on $f(t)'$, and will go to zero once $X(t)$ has replaced the other variant, i.e. the estimator recalibrates itself. We can now bound the bias

$$R^{w_2}(t) - R(t) \le \gamma^{-1}\max_t \frac{(c-1)f(t)'}{1+(c-1)f(t)} \le \gamma^{-1}\frac{(c-1)\max_t f(t)'}{1+(c-1)\min_t f(t)} \quad (49)$$

Here we take $\max_t f(t)' = \max_t \phi' f(t)(1 - f(t)) = \phi'/4 = s\gamma/4$, and we bound $\min_t f(t) = 0$. This yields the bound

$$R^{w_2}(t) - R(t) \le \frac{(c-1)s}{4} \quad (50)$$

#### (iii) Estimates are biased by arbitrary changes in shedding load distribution

We now study what happens when we wrongly assume that $g_x = g_y$, while in fact the shape of the shedding load profiles are different. Following Equation 1 we have that:

$$\left(I * g^{-1}\right)(t) \approx l^{-1}I(t + \mu) \Rightarrow \left(X * g_x * g_y^{-1}\right)(t) \approx \frac{l_x}{l_y}X(t + \Delta_\mu) \quad (51)$$

Where $\Delta_\mu = \mu_y - \mu_x$. For simplicity we will assume here that $l_x = l_y$. We then have

$$X(t + \Delta_\mu) \approx X(t) + X'(t)\Delta_\mu \approx X(t)\left(1 + (\beta_x - \gamma)\Delta_\mu\right) \quad (52)$$

Following above, and assuming a constant $\beta_x$, we find the approximate bias for our estimator $R^{w_2}(t)$ along with upper bound

$$R^{w_2}(t) - R(t) \approx \gamma^{-1} \frac{(\beta_x - \gamma)\Delta_\mu f(t)'}{1 + (\beta_x - \gamma)\Delta_\mu f(t)} \leq \frac{s(\beta_x - \gamma)\Delta_\mu}{4} \quad (53)$$

If we drop the assumption of constant $\beta_x$, we can further find an approximate bias term for our estimator $R^{w_2}(t)$ and an upper bound

$$R^{w_2}(t) - R(t) \approx \gamma^{-1} \frac{(\beta_x(t) - \gamma)\Delta_\mu f(t)' + \Delta_\mu \beta_x(t)' f(t)}{1 + (\beta_x - \gamma)\Delta_\mu f(t)} \leq \frac{s(\beta_x(t) - \gamma)\Delta_\mu}{4} + \Delta_\mu \beta_x(t)' \quad (54)$$

#### Dispersion of the generation interval time

In the derivations of closed-form expressions for the biases in selection advantage and *Re* due to reduced or increased shedding, we assumed an exponentially-distributed generation time interval. We simulated data to assess the impact of underdispersed or overdispersed generation time intervals on the estimates of $R_e$ and selection advantage. We simulated two distinct time series of SARS-CoV-2 infections, each for a different variant: the first variant started from 1500 cases with a constant $R_e$ of 0.6, and the second started from a single case with a constant $R_e$ of 2.2. The simulations largely followed the simulation framework used in Huisman et al.[2,20]. Infections were simulated forward in time using the renewal equation framework described by Cori et al[2120,21]. We relaxed the exponentially-distributed generation time to a Gamma-distributed generation time, with constant mean but varying variance. The overall $R_e$ was then estimated from the simulated infection incidence time series, once considering the two time series untouched and once having the second time series scaled down by 50% to simulate undershedding. The $R_e$ values were computed using the package *EpiEstim*[18].
